# From Web to RheumaLpack: Creating a Linguistic Corpus for Exploitation and Knowledge Discovery in Rheumatology

**DOI:** 10.1101/2024.04.26.24306269

**Authors:** Alfredo Madrid-García, Beatriz Merino-Barbancho, Dalifer Freites-Núñez, Luis Rodríguez-Rodríguez, Ernestina Menasalvas-Ruíz, Alejandro Rodríguez-González, Anselmo Peñas

## Abstract

This study introduces *RheumaLinguisticpack* (*RheumaLpack*), the first specialised linguistic web corpus designed for the field of musculoskeletal disorders. By combining web mining (i.e., web scraping) and natural language processing (NLP) techniques, as well as clinical expertise, *RheumaLpack* systematically captures and curates structured and unstructured data across a spectrum of web sources including clinical trials registers (i.e., ClinicalTrials.gov), bibliographic databases (i.e., PubMed), medical agencies (i.e. EMA), social media (i.e., Reddit), and accredited health websites (i.e., MedlinePlus, Harvard Health Publishing, and Cleveland Clinic). Given the complexity of rheumatic and musculoskeletal diseases (RMDs) and their significant impact on quality of life, this resource can be proposed as a useful tool to train algorithms that could mitigate the diseases’ effects. Therefore, the corpus aims to improve the training of artificial intelligence (AI) algorithms and facilitate knowledge discovery in RMDs. The development of *RheumaLpack* involved a systematic six-step methodology covering data identification, characterisation, selection, collection, processing, and corpus description. The result is a non-annotated, monolingual, and dynamic corpus, featuring almost 3 million records spanning from 2000 to 2023. *RheumaLpack* represents a pioneering contribution to rheumatology research, providing a useful resource for the development of advanced AI and NLP applications. This corpus highlights the value of web data to address the challenges posed by musculoskeletal diseases, illustrating the corpus’s potential to improve research and treatment paradigms in rheumatology. Finally, the methodology shown can be replicated to obtain data from other medical specialities. The code and details on how to build *RheumaL*(*inguistic*)*pack* are also provided to facilitate the dissemination of such resource.

## 1 Introduction

Most of the existing data worldwide are unstructured Tam Harbert [2021]. Reports estimate that these data comprise about 80 to 90 percent of all newly generated data Forbes Tech Council [2017]. In medicine, about 80% of total electronic health record data is unstructured Li et al. [2022]. Therefore, in recent years, there has been a growing interest in the application of natural language processing (NLP) techniques within the clinical field Wang et al. [2020], to structure and use these data. The adoption of advanced deep learning (DL) models, including transformer-based architectures Vaswani et al. [2017], such as bidirectional encoder representations from transformers (BERT) Devlin et al. [2019] or other large language models (LLM), has contributed significantly to this growth of interest.

NLP techniques are commonly used to gain insights and uncover relevant non-previously exploited information. Information extraction, retrieval, or knowledge discovery are some of the fields within NLP that try to transform unstructured data into actionable knowledge. However, like any other artificial intelligence algorithm, NLP approaches need data to build robust systems capable of addressing current challenges Khurana et al. [2023], Zhou et al. [2020]. In medicine and clinical research, highly relevant information can be found and extracted from the Web to form a corpus that can be used to train those NLP systems. To begin with, there are multiple websites linked to databases and search engines where information on clinical trials can be found, such as the Cochrane Controlled Trials Register (CENTRAL), ClinicalTrials.gov, or EU Clinical Trials Register. Scientific abstracts and article-related information can also be retrieved from bibliographic databases such as PubMed, Scopus, or Embase; whereas drug leaflets can be found on governments and regulatory agencies’ websites, such as European Medicines Agency (EMA). Moreover, social media and forums may contain relevant clinical information that could improve NLP algorithms performance. These last data sources have received special attention in recent years in the healthcare sector Nawaz et al. [2017]. In fact, patients are prone to use social media websites for health-related purposes Studenic et al. [2018], Taik et al. [2024], Blackie et al. [2023], and to express their feelings Wilson et al. [2023], Abbasi-Perez et al. [2023], creating what is known as patient-generated data. Finally, accredited health websites provide trustworthy health information which can complement the previous data sources for training specialised AI systems.

In the field of rheumatic and musculoskeletal diseases (RMDs), the application of AI, NLP, and LLMs is not new Madrid-Garcia et al. [2023a,b]. In fact, NLP approaches have been used in the past to answer different RMDs research questions Jorge et al. [2019], Maarseveen et al. [2020], Humbert-Droz et al. [2023], Ivorra et al. [2024]. However, none of these efforts have focused on developing a domain-specific corpus that captures the unique language and terminology inherent to the field of RMDs, which could significantly benefit future research studies. Having a domain-specific corpus would boost NLP research in rheumatology, facilitating more accurate model training, improving the granularity of NLP applications, creating new research avenues, and enabling more nuanced understanding and analysis of RMD-related texts. With such a corpus, new research lines could blossom, such as chatbots fine-tuned with RMDs specific knowledge or embeddings creation.

Multiple efforts have been made to build medical corpora in clinical research. However, many of these corpora are designed to address specific tasks and are not often considered reusable resources for other researchers, resulting in them not being shared. In addition, the corpus creation process is not often described in depth and is relegated to the background, so it can not be replicated. Given these challenges, the objective of this study is to illustrate how rheumatology-related medical web data can be extracted from multiple sources using web mining approaches, among others, to build a domain-specific corpus that can be useful in a multitude of scenarios.

## 2 Related work

In recent years, researchers have developed corpora using a wide variety of data sources. For instance, authors in Kury et al. [2020], presented Chia, a corpus obtained from ClinicalTrials.gov with 1,000 actively recruiting, interventional, phase 4 studies; and made it publicly available. The objective with Chia was to build the first annotated corpus for clinical trial eligibility criteria, that could be used for training machine learning systems for information extraction, or electronic phenotyping.

Researchers in Collins et al. [2024] presented a biomedical corpus containing oncology information using PubMed abstracts. For this purpose, the authors focused on eight PubMed cancer-related journals and downloaded the first 100 abstracts from each journal. A total of 800 abstracts were stored in individual text files and processed to include only the abstract title and body. The objective of this corpus was to train systems capable of extracting the most important details from cancer genomics experiments. In Wang et al. [2021], authors used more than 100,000 PubMed abstracts to generate ophthalmology-specific word embeddings. Then, they used the embeddings to predict visual prognosis Gui et al. [2022]. On their behalf, the researchers in Beam et al. [2019] used 1.7 million full-text journal articles from PubMed to generate *cui2vec* embeddings. These embeddings were later used to identify functional relations among diseases Bugrim [2023].

The authors in Foufi et al. [2019], built a corpus using social media data (i.e., Reddit) with more than 17k posts comprising chronic diseases mentions from 19 subreddits (e.g., r/cancer, r/MultipleSclerosis, r/rheumatoid, r/testicularcancer). These comments were merged into a single dataset. Crawlers, software that facilitates the recursive process of discovering and downloading web pages by following links extracted (or harvested) from already known sites, were used to collect the data. Reddit, was also employed in Okon et al. [2020]. In this study, the authors focused on the most common skin diseases worldwide identified by the Global Burden of Diseases to search and collect data from 176,000 Reddit comments.

Medical agencies such as EMA have been used in the past to build, LeMe-PT, a portuguese corpus capable of generating competitive semantic models Simões and Gamallo [2021]. Another Spanish, corpus that included leaflets obtained from the Spanish medical agency (i.e., CIMA) was constructed in Campillos Llanos et al. [2022]. This corpus was intended for text simplification.

Data from MedlinePlus, have also been extracted for different purposes. In Denecke and Nejdl [2009], the authors crawled 750 pages from MedlinePlus and other sources to give an overview of content differences in the various social media resources on health-related topics. In Segura-Bedmar et al. [2016], scholars also used MedlinePlus to build a Spanish corpus of drug leaflets and diseases.

## 3 Methodology

A six-step methodology was applied to build *RheumaLpack* corpus. This methodology was based on expert guidance from a rheumatologist of the Hospital Clínico San Carlos, Madrid, Spain. The steps followed are shown in Figure 1.

**Figure 1:**
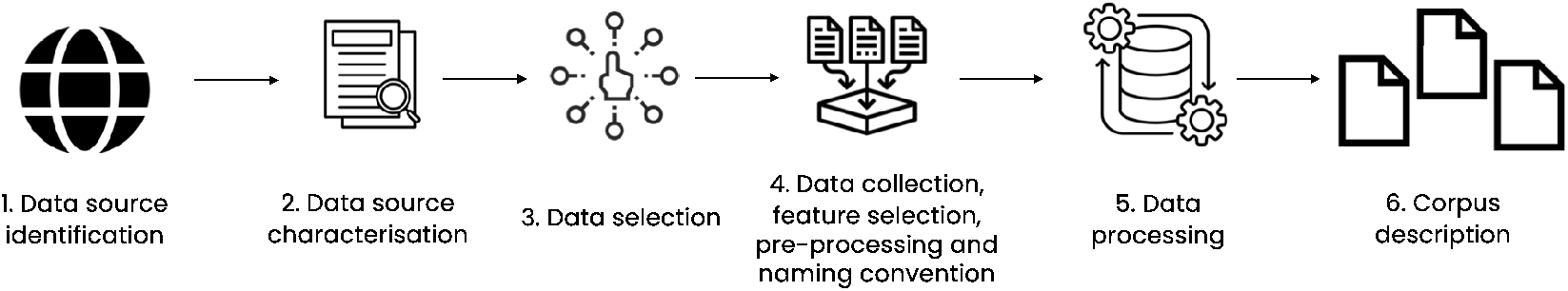
Methodology followed for building *RheumaLpack corpus*. Author’s elaboration

1. **Data source identification**: a set of eleven criteria was proposed to select the different data sources that finally comprised *RheumaLpack*. The sources were selected by consensus, after considering the criteria shown in Table 1. Variability criterion was the most important one, as it was agreed that at least one clinical trial registry, bibliographic database, medical agency, social media website and accredited health website had to be used.
2. **Data source characterisation**: after identifying all relevant data sources, a brief description of them was provided.
3. **Data selection**: considering the potential data volume and diversity, only relevant data in the context of RMDs was selected for collection. To determine the data to be extracted from clinical trials registries or bibliographic databases, search engines queries using RMDs related keywords such as “musculoskeletal diseases” or “rheumatic diseases” were conducted. In addition, to meet with the *timeliness* and *relevance* criteria, only data from January 1^st^, 2000, to December 31^th^, 2023 were included. To comply with the *accuracy* criterion, only information from non-withdrawn medications was collected. The decision on which medications to include was made based on the judgment of a rheumatologist, the recommendations of rheumatology scientific societies (i.e., Sociedad Española de Reumatología (SER)) or the characteristics of the drug itself (i.e., widely used RMDs drugs, such as, disease-modifying antirheumatic drugs (DMARDs)). For social media data, we prioritised active sites, this is, those with daily to annual activity. Finally, data from accredited health websites was manually selected by a rheumatologist.
4. **Data collection, feature selection, pre-processing and naming convention**: data collection was carried out according to the *accessibility* criterion. There were two primary methods for data extraction:
  - Data accessible via API, REST-API interfaces or API wrappers: this was the default option. Prior to data extraction through APIs, a feature selection step was done. This process was essential for specifying the exact dataset parameters to be queried from the API.
  - Data not accessible through API interfaces:
    - Data available through direct downloads: when the data source provided ways to download the data directly, without coding, this option was considered the default option.
    - Data not available through direct downloads: web scraping techniques using Python packages such as [Beautiful Soup](https://pypi.org/project/beautifulsoup4/) were used. This stage also included data pre-processing (e.g., removal of escape characters such as newlines or tabulations). Information from the same data source could be extracted in batches of different sizes, if it facilitated data collection. Moreover, for practicality, the information gathered from each data source could be stored in individual files or in larger data structures with each line representing a single record. This approach made data management and retrieval easier. Unique naming conventions were established to ensure a distinct identifier for each file within *RheumaLpack*. Data collection extended beyond unstructured fields to include structured fields that could provide additional value to researchers. Eventually, some health websites implemented paywalls to certain pages or antiscraping techniques. Therefore, websites on a paywall were not scraped. To avoid antiscraping techniques, customisation of the *user agent string* was considered.
  - **Data processing**: to ensure the versatility of the corpus in various domains, only minimal generic processing was planned. This was designed to create a corpus that was as broadly applicable as possible, enabling its use across a wide range of tasks. Depending on how the data were obtained, the processing varied. For instance, for data obtained with web scraping techniques, the pre-processing was done at the same time of data collection, and no further processing was conducted. For the rest of the data, duplicates were managed; and the data from the same source extracted in different batches were combined. Following the *completeness* criterion, records that did not contain the expected data (e.g., articles without abstracts) were removed. Following the *non-personal data* criterion, personally identifiable information was also managed in this step.
  - **Corpus description**: upon completion of the preceding steps, the corpus built (i.e., *RheumaLpack*) is described, detailing its characteristics (e.g., number of records per source, size, hierarchy).

**Table 1:** Data source identification criteria.

| Criteria | Description |
| --- | --- |
| Variability | Variability across three dimensions was promoted: a) Expert vs. Non-expert information b) Type of data, c) Coverage of different aspects of RMDs (e.g., diseases, symptoms, treatments) |
| Accessibility | Ease of obtaining data, already developed tools to extract data (e.g., REST-API, direct downloads, scraping techniques) |
| Renowned | Data sources should be widely recognised and respected worldwide to enhance the legitimacy, comprehensiveness, and acceptance of the research findings derived from the corpus |
| Timeliness | Data sources should encompass both contemporary information and historical data |
| Non-personal data by default | Data sources devoid of information governed by the GDPR were prioritized to reduce the risk of unintentionally breaching individuals' privacy rights |
| Relevance | Meaningful information and applicable to the research questions and objectives concerning RMDs |
| Ease of processing and analysis | Data sources should contain data that are easily manageable and processable by analytical tools |
| Language coverage | Data sources should contain data in English |
| Geographical coverage | Data source should cover different geographic locations or populations |
| Completeness | Data from the different data sources should be as complete and error-free as possible |
| Accuracy | Data should be correct, precise, and reflective of the current state of knowledge regarding RMDs |

## 4 Results

### 4.1 Data source identification

Fourteen different sources were initially evaluated, see Supplementary Material Text and Supplementary Table 1. After consensus, and considering the criteria shown in Table 1, seven of them were selected: *ClinicalTrials, PubMed, European Medicines Agency, Reddit, MedlinePlus, Harvard Health Publishing School*, and *Cleveland Clinic*. Before finalising the selection and determining the suitability of each data source for inclusion in *RheumaLpack* corpus, preliminary tests to extract information from that source were conducted. This step was essential in evaluating the practicality and efficiency of retrieving data from each potential data source.

### 4.2 Data source characterisation

#### ClinicalTrials.gov

ClinicalTrials.gov, the world’s largest database of clinical trials funded both privately and publicly, is integrated into the International Clinical Trials Registry Platform. This online platform offers open-access to its repository, which contains over 490,000 registered studies, of which 54% are conducted outside the US, with an annual growth per year of 35.000-40.000 studies.

#### PubMed

PubMed is a freely accessible search engine owned by the National Library of Medicine which allows accessing primarily the MEDLINE database. PubMed indexes articles from more than 50,000 journals and the number of papers indexed per year exceeds one million. Until 2023 it contained more than 36 million citations and abstracts of biomedical literature.

#### EMA

According to its official webpage, EMA is a decentralised agency of the European Union responsible for scientific evaluation, supervision, and safety monitoring of medicines. EMA publishes clear and impartial information about medicines and their approved uses.

#### Reddit

Reddit is among the most prominent social platforms on the web Proferes et al. [2021]. As of October 2023, it hosts over 100,000 active communities and receives daily visits from more than 70 million users, making it one of the top 20 most visited sites in the world Reddit, Inc. [2023]. Discussions on Reddit are primarily public, and different RMDs-related communities exist. Data from these subreddits, can be considered patient-generated data.

#### Accredited health websites

MedlinePlus is a digital platform, provided by the National Library of Medicine, that provides a wealth of information on health topics, including diseases and drugs. It sources content from approximately 500 selected organizations and offers nearly 22,000 links to authoritative health information in English Harvard Health Publishing offers a comprehensive encyclopedia featuring over 500 diseases and conditions, authored by the faculty of Harvard Medical School. Additionally, Cleveland Clinic is a leading medical organisation that provides not only healthcare services but also hosts extensive online libraries of medical resources.

### 4.3 Data selection

#### ClinicalTrials.gov

Clinical trials retrieved from ClinicalTrials.gov, after filtering by the *Condition/disease*: *Rheumatic Diseases* and published through January, 1^st^, 2000 to December 31^st^ 2023 were selected; and the clinical trial number (NCT) identified, n = 9,144. Supplementary Figure 1 illustrates the trend in the publication of clinical trials concerning RMDs over time.

#### PubMed

Only abstracts that were indexed in MEDLINE PubMed through January, 1^st^, 2000 to December, 31^st^ 2023 were selected. To narrow the search to scientific journals specialised in rheumatology, the JCR index was used. Hence, the journals classified by JCR as “RHEUMATOLOGY SCIE” were identified, see Supplementary Table 2, and we proceeded to identify the PubMed identifiers (PMID). For that purpose, manual queries with the name of each journal were run in PubMed adding “[Journal]” at the end (e.g., “Annals of the rheumatic diseases”[Journal]). It should be noted that PubMed limits the download size to 10,000 items, so multiple queries could be run in case a journal had more than 10,000 publications during the study period. A total of 122,426 PMIDs were recovered.

#### EMA

The EMA provides a Public Excel sheet, *European public assessment reports* (*EPARs*) *for human and veterinary medicines*, specifying the therapeutic area of application of medicines that have been granted or refused a marketing authorisation. From this excel, all the URLs of rheumatology approved drugs were collected, n = 44. Each of the identified URLs contains the *product information* of each drug, this is, a PDF file containing a summary of the product characteristics, conditions, or restrictions regarding supply and use; among others.

#### Reddit

Since Reddit was founded in 2005, only data from that date to 2023 were selected. The subreddits related to RMDs were identified using the Reddit search engine, after filtering by *Community*. A list of common diseases and words employed in rheumatology was used for this purpose (i.e., arthritis, autoimmune, back pain, backpain, behcet, behcets, fibromyalgia, gout, lupus, myositis, psoriasis, raynaud, raynauds, rheumatology, scleroderma, sjogren, sjogrens, spondylitis, tendinitis, thritis, uveitis, vasculitis). In Supplementary Table 3 the complete list of identified communities can be seen. Only RMDs-related subreddits, assessed by a rheumatologist, within the 40,000 most active subreddits worldwide, according to pushshift.io, were selected, n = 12, namely r/ankylosingspondylitis, r/Autoimmune, r/autoimmunity, r/backpain, r/Fibromyalgia, r/gout, r/lupus, r/PsoriaticArthritis, r/rheumatoid, r/rheumatoidarthritis, r/Sjogrens, and r/Thritis.

#### Accredited health websites

Since accredited health websites contain information on all medical specialities, a manual assessment of the topics related to RMDs was conducted by a rheumatologist. To facilitate this task, a Python script which extracted the name of each topic and its URL was developed and presented to the physician. Finally, this physician decided what information should be further extracted. The total number of selected items to retrieve from MedlinePlus, Harvard Health Publishing, and Cleveland Clinic was 282, 47 and 320 respectively. The outcome of this step was a collection of URLs designated for scraping.

### 4.4 Data collection, feature selection, pre-processing and naming convention

#### ClinicalTrials.gov

ClinicalTrials.gov provides a REST-API that can be used to extract all the information related to clinical trials. API calls were made directly through Python scripting, filtering by *Rheumatic Diseases* condition/disease. Twenty-six variables were collected after specialist advice: *NCTId, Official Title, Brief Title, Overall Status, Study Start Date, Completion Date, Study Type, Conditions, Keywords, Brief Summary, Detailed Description, Eligibility Criteria, Sex, Minimum Age, Study Population, Lead Sponsor, Responsible Party Investigator Full Name, Investigator Full Name, Primary Outcomes, Secondary Outcomes, Has Results, Organization Full Name, Phases, Enrollment Count, Allocation, Intervention Model*. The textual most relevant ones were *Brief Summary and Detailed Description*. Since the ClinicalTrials.gov API limits the retrieval to 1,000 clinical trials per request, multiple queries were made. All the initially selected clinical trials, n = 9,144 were gathered.

#### PubMed

R’s *rentrez* library Winter [2017] was used to collect the following information from each article *PMID, DOI, MeSH keywords, volume, issue, pages, abstract, has abstract, publication type, language, PubMed central papers citation, sort first author and affiliation*. These data were gathered in three batches: the first batch containing the abstract and other publication details such a MeSH keywords; the second batch containing the language, the publication type and the PubMed central papers citation; and the third one containing affiliation data.

#### EMA

A Python script was built to download the *Product information* PDFs from each identified drug from EMA website, and to convert the PDF files into .txt files. This last step was done with the pypdf package. Afterwards, basic preprocessing was performed, including removing special characters (e.g., \n, \r and \t) and collapsing multiple spaces. All the initially selected drug leaflets, n = 44, were gathered.

#### Reddit

Pushshift.io Baumgartner et al. [2020], stuck_in_the_matrix, Baumgartner [2024] was used to extract subreddits data (i.e., *submissions* and *comments*) from the 12 selected subreddits. A publicly accessible torrent from [Academic Torrents](https://academictorrents.com/details/56aa49f9653ba545f48df2e33679f014d2829c10) was employed to obtain such data. After downloading, a public script accessible via GitHub Watchful1 [2024] was used to decompress the files, which were stored in .zst format. During this decompression phase, the variables to be extracted were specified. For *submissions*, these included *id, author, title, created utc, and selftext*. Similarly, for *comments*, the same information was collected along with the *parent id* (i.e., the submission/starting post id associated with each comment). Some additional parameters that can be included during the extraction phase are shown in Supplementary Table 4.

#### Accredited health websites

Different Python scripts were developed for the different accredited health websites. Each script uses the Beautiful Soup Python package to scrape and preprocess the data. Only the relevant textual information contained on each website was downloaded, determined by a rheumatologist. Therefore, manual rules specifying the beginning and ending text had to be defined. Moreover, since different health categories within the same health website can be displayed differently, more than one script per health website was written when needed. Not all the identified items in the previous steps were retrieved since 13 of them were behind a paywall and therefore skipped.

### 4.5 Data processing

#### PubMed

Not all the selected articles had an abstract, since this information is only collected for a certain type of articles (e.g., original research articles, reviews), therefore we excluded those without this information. A total of 96,004 abstracts were finally gathered. The first two batches were merged into a single document, while the third batch, pertaining to affiliations, was kept separate. A detailed list with the number of articles with abstract per journal and by year can be seen in Supplementary Table 5.

#### Reddit

A Python script was built to combine the *comments* and *submissions* information of each subreddit into a single file. In addition, the information was sorted by id, parent id and date of submission to ensure that the initial posts and their corresponding comments were presented in a sequential manner. After that, and to preserve privacy Benton et al. [2017], the id values that uniquely identify a post/comment on Reddit were encrypted using the MD5 hashing algorithm, the authors’ nicknames were deleted; and a time of 180 was added/subtracted to the creation date of the post/comment to anonymise the dataset. This was done by grouping by id, to ensure that all comments on the same topic had the same time shift. Eventually, 12 files and five columns remained: *hashed id* with MD5 algorithm, *anonymised date of submission in UTC milliseconds, post title, body / text message, and row type* (i.e., post/submission). The total number of posts and submissions for each subreddit can be seen in the Supplementary Table 6.

### 4.6 Corpus description

The *RheumaLpack* corpus comprises nearly three million records sourced from seven distinct data sources, with a total size of 1.34 GB. Although originally conceived as a linguistic resource, *RheumaLpack* also includes structured data from ClinicalTrials.gov and PubMed. Some clinical studies have shown that the combination of both types of data results in algorithms with higher predictive power Zhang et al. [2020]. However, the most important variables in these datasets are textual (e.g., *Brief Summary and Detailed Description* for ClinicalTrials.gov, *Abstract* for PubMed, and *title and body* for Reddit). Table 2 shows the main characteristics of *RheumaLpack*. This corpus could be classified as a monolingual, non-annotated, and dynamic resource since it only contains data in English, has not been processed with annotations, and new information is expected to be gathered yearly by subtly modifying the scripts.

**Table 2:** RheumaLPack corpus documents characteristics.

| Source | Subset | N of documents | N of registers | Data | Size |
| --- | --- | --- | --- | --- | --- |
| ClinicalTrials.gov |  | 1 | 9,144 | Study protocol, brief summary, detailed description | 37 MB |
| PubMed | Abstracts | 1 | 96,004 | Abstract, journal, title, first author | 205 MB |
|  | Affiliation | 1 | 670,843 | Affiliation centre, contact details | 80.3 MB |
| European Medicines Agency |  | 44 |  | Drug leaflets | 14 MB |
| Reddit |  | 12 | 2,269,311 | Social media data, patient generated data comments and posts | 1 GB |
| MedlinePlus | Drugs | 135 |  | Websites with information on drugs, diseases, symptoms, tests, procedures | 1.5 MB |
|  | Others | 147 |  |  | 0.5 MB |
| Harvard Health Publishing |  | 34 |  |  | 0.2 MB |
| Cleveland Clinic | Diseases & Conditions | 178 |  |  | 1.7 MB |
|  | Treatments | 41 |  |  | 0.3 MB |
|  | Diagnostics & Testing | 60 |  |  | 0.46 MB |
|  | Symptoms | 41 |  |  | 0.3 MB |

Supplementary Excel File “Data ID” includes the following information: clinical trial numbers, abstract PMIDs, URLs for drug leaflets; and URLs of accredited health websites from which data was collected. All the Reddit data contained in the torrent described earlier was used.

## 5 Discussion

To our knowledge, no previous attempts have been made to create an accessible corpus that combines data from clinical registries (e.g., ClinicalTrials.gov), bibliographic databases (i.e., PubMed), medical agencies (i.e., EMA), social media (i.e., Reddit), accredited health websites (i.e., MedlinePlus, Harvard Health Publishing, and Cleveland Clinic), in medicine and specifically in RMDs. One of the strengths of this study is that it presents a methodology that can be replicated in other medical specialities. In addition, the developed code is accessible on GitHub with instructions on how to run it, facilitating its use by researchers and the scientific community.

Some of the identified potential uses of *RheumaLpack* could be:

1. AI systems training:
  - Predictive systems: *RheumaLpack* could be used to train systems for named-entity recognition, relation extraction, sentence classification, to study the polarity of a text, or to create embeddings; among others.
  - Generative systems: *RheumaLpack* could be used to integrate rheumatology specific knowledge to open source LLMs Labrak et al. [2024], such as Llama 3. Once done, these models could be deployed in a specialised clinic to help the junior rheumatologist summarise the clinical care plan of a report, or could be used as a chatbot for patients seeking answers to their questions after office hours. For instance, authors in Jia et al. [2024], developed OncoGPT, this was done by fine-tuning Llama with oncology-related conversations.
2. Evaluation frameworks, benchmarking, gold standards creation and shared tasks: the corpus presented could be used to evaluate the performance of novel AI systems or could be used in shared tasks and challenges within the research community.
3. Research: *RheumaLpack* could enable new studies in NLP, data mining, information retrieval, machine translation, and machine learning applied to the rheumatology domain. Researchers could take advantage of the diverse data sources to investigate topics such as:
  - Drug safety and efficacy analysis: by analyzing patient discussions on social media and information from drug leaflets, researchers can gain insights into real-world drug safety and efficacy. This could complement traditional clinical trial data, providing a more holistic view of treatment outcomes.
  - Patient education and engagement strategies: understanding the types of questions and concerns patients have, as reflected in social media data, can inform the development of more effective patient education materials and engagement strategies. This could lead to improved patient adherence and outcomes.

### 5.1 Limitations

While this study has provided important insights, it is not without some limitations:

- Information extracted from Reddit may not accurately represent RMDs patients, who are predominantly elderly and female. In contrast, Reddit’s user base is mainly (58%) young males aged 18 to 34, as noted in Proferes et al. [2021].
- Texts from abstracts or drug leaflets, which use technical and formal language, differ significantly from the colloquial and often ironic or sarcastic language found on social media platforms like Reddit. Combining both data types may reduce the performance of AI solutions.
- PubMed abstracts could have been obtained using MeSH terms in the searches: such as “rheumatology”[MeSH Terms]. In this way, we would not be dependent on the JCR classification.
- Other data sources could have been included in *RheumaLpack*, such as Wikipedia webpages or anonymised electronic medical records. Moreover, open access full-text rheumatology articles could be downloaded and converted into text files, as we did with EMA product information files. This would enrich the corpus with more research data.
- The scripts designed to scrape data are highly dependent on the specific layout of the websites from which they gather information. Even minor modifications to the website’s design can disrupt the functionality of these scripts. Rather than creating individual scripts for each website, a single script that downloads all textual content and subsequently processes it could be developed.
- The legality of scraping has always been questioned. In fact, the use of scraping techniques to reproduce and redistribute data can lead to copyright infringements. This has been widely discussed in the literature Krotov et al. [2020], Gold and Latonero [2017]. In our case, we would like to point out:
  - No personal data are extracted
  - There is no financial gain/compensation
  - The data extracted are obtained from public sources to which everyone has access to them
  - Ownership of extracted data is recognised
  - Efforts are made to use the server responsibly by minimizing the number of data extraction calls
  - In Spain, where this study has been conducted, there is no specific law prohibiting web scraping
  - No data are shared, only the code to obtain it.
- Drug leaflets are available in various languages; in fact, EMA offers this information in more than 20 languages. In this work we focused on English documents, however as showed, drug leaflets information in other languages could have been extracted easily obtained to build a parallel multilingual drug leaflets corpus.
- The data typically employed for training or fine-tuning LLMs is conversational in nature. However, the *RheumaLpack* dataset does not contain this type of data due to its limited availability.

## 6 Conclusion

In this study, we have shown how web-accessible data can be used to build a multipurpose specialised medical corpus, called *RheumaLpack*. To create this resource, we employed a six-step methodology and combined different approaches to obtain the data (i.e., REST-API, scraping, direct downloads). The data sources used in this study include: clinical registries (e.g., ClinicalTrials.gov), bibliographic databases (i.e., PubMed), medical agencies (i.e., EMA), social media (i.e., Reddit), and accredited health websites (i.e., MedlinePlus, Harvard Health Publishing, and Cleveland Clinic). This corpus comprises nearly three million data points that cover clinical information, research, and social media data. In fact, we have established the foundation for the construction of specialised corpora in rheumatology. This may inspire the scientific community within this medical field to boost this research line, taking advantage of recent interest in LLMs. Finally, this work seeks to make rheumatology-related data more accessible and analyzable, opening new paths for groundbreaking research and promising advances in understanding and addressing rheumatic diseases with NLP techniques. The code and details on how to build *RheumaL*(*inguistic*)*pack* are also provided on request to facilitate the dissemination of such resource.

## Data Availability

Raw data from RheumaLpack is not available as these data may be protected by copyright. However, all the code developed for building the corpus, along with instructions on how to use it, is provided on request.

## Data availability statement

Raw data from *RheumaLpack* is not available as these data may be protected by copyright. However, all the code developed for building the corpus, along with instructions on how to use it, is provided on request. Access is not guaranteed as the webpages may change, and therefore the scraping algorithms become useless. In addition, API access requirements may change over time. The corresponding author will be happy to provide further indications to researchers interested in deploying the algorithms to build *RheumaLpack* locally.

## CRediT author statement

**Alfredo Madrid-García**: Conceptualization of this study, methodology, coding, review, writing (original draft preparation). **Beatriz Merino-Barbancho**: Methodology, review, coding. **Dalifer Freites-Núñez**: Methodology, writing. **Luis Rodríguez-Rodríguez**: Methodology, review. **Ernestina Menasalvas-Ruíz**: Methodology, review. **Alejandro Rodríguez-González**: Methodology, review. **Anselmo Peñas**: Conceptualization of this study, methodology, review.

All of the authors were involved in the drafting and/or revising and/ or publishing the manuscript.

## Supplementary Material Files

- Supplementary Excel File Data ID: Excel file that includes the identifiers of the clinical trials, PubMed abstracts and the URLs of the websites downloaded.
- Code to generate RheumaLpack

## Acknowledgement

The authors would like to thank the *pushshift*.*io* team, specially to Watchful1 Reddit user.

## Supplementary Material

### Supplementary Text

#### Data source identification

Initially, fourteen different sources were considered: [European Medicines Agency (EMA)](https://www.ema.europa.eu/en/homepage), [Centers for Disease Control and Prevention](https://www.cdc.gov/health-topics.html), [Centro de Información de Medicamentos (CIMA)](https://cima.aemps.es/cima/publico/home.html), [Cleveland Clinic](https://my.clevelandclinic.org/health/diseases), ClinicalTrials.gov, [Harvard Health Publishing](https://www.health.harvard.edu/health-a-to-z), [Johns Hopkins Medicine](https://www.hopkinsmedicine.org/health/conditions-and-diseases/), [Mayo Clinic](https://www.mayoclinic.org/diseases-conditions), [MedlinePlus](https://medlineplus.gov/), [Mount Sinai](https://www.mountsinai.org/health-library), [PubMed](https://pubmed.ncbi.nlm.nih.gov/), [Reddit](https://www.reddit.com/?rdt=35225), [WebMD](https://www.webmd.com/), and Wikipedia.

After expert discussion, the data sources that best matched the criteria were ClinicalTrials.gov, PubMed, EMA, Reddit, MedlinePlus, Harvard Health Publishing, and Cleveland Clinic. It was not necessary for them to strictly meet all of the aforementioned criteria. For instance, in the case of Reddit, some of the data shared by users on posts could be considered personal data under the GDPR, especially if these data can be used to identify a specific individual, either on its own or in combination with other information. On the basis of the variability criterion, expert sources such as PubMed, EMA, MedlinePlus, or ClinicalTrials.gov were chosen for their authoritative and comprehensive data on clinical research findings, treatment protocols, and drug information. Conversely, Reddit was selected as a non-expert source to capture patient opinions, experiences, and discussions, providing valuable insights into the patient perspective. This diverse selection allowed the corpus to include a wide range of data types, from social media content to scholarly articles and clinical trials information, ensuring a holistic view of diseases from multiple angles: pharmacological treatments, patient viewpoints, and research advances.

### Supplementary Tables

**Supplementary Table 1:**
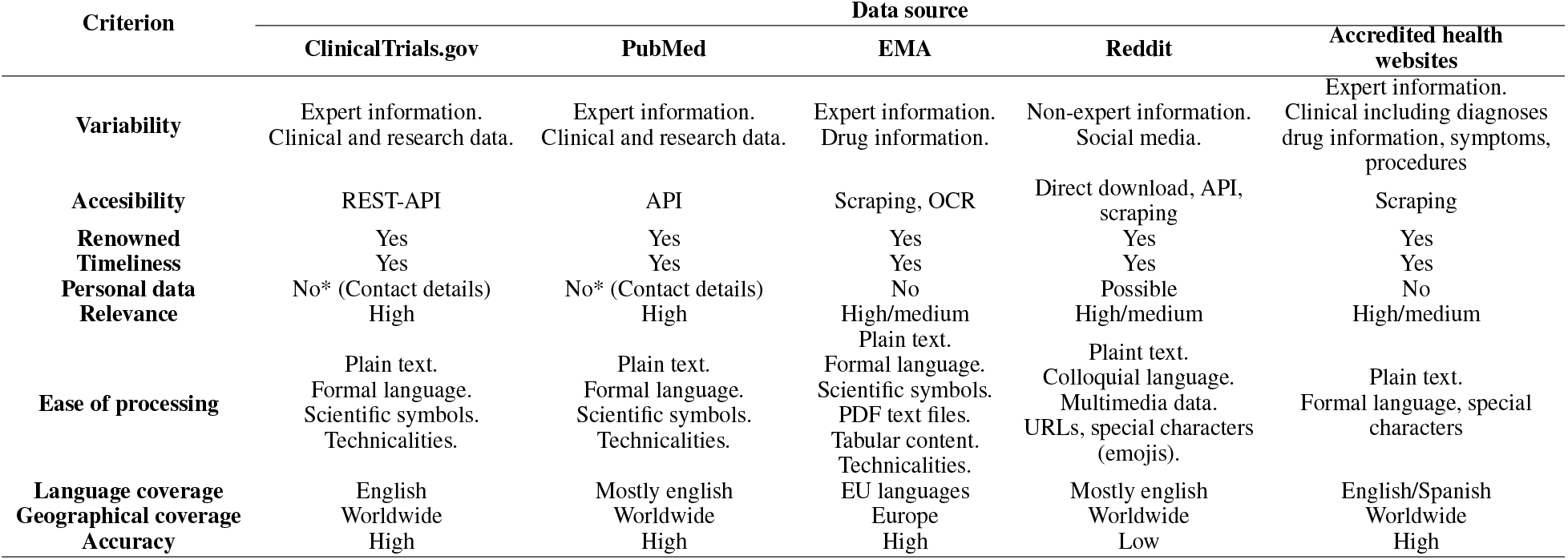
Criteria evaluation for the selected data sources

**Supplementary Table 2:** Rheumatology journals classified by JCR as “RHEUMATOLOGY SCIE”. The journal name is written as appears in JCR webpage

| Journal name |  |
| --- | --- |
| Nature Reviews Rheumatology | Rheumatology and Therapy |
| ANNALS OF THE RHEUMATIC DISEASES | CLINICAL AND EXPERIMENTAL RHEUMATOLOGY |
| Lancet Rheumatology | CLINICAL RHEUMATOLOGY |
| Arthritis & Rheumatology | JCR-JOURNAL OF CLINICAL RHEUMATOLOGY |
| Osteoarthritis and Cartilage | LUPUS |
| RMD Open | Pediatric Rheumatology |
| RHEUMATOLOGY | International Journal of Rheumatic Diseases |
| BEST PRACTICE & RESEARCH IN CLINICAL RHEUMATOLOGY | BMC MUSCULOSKELETAL DISORDERS |
| CURRENT OPINION IN RHEUMATOLOGY | Advances in Rheumatology |
| Current Rheumatology Reports | RHEUMATIC DISEASE CLINICS OF NORTH AMERICA |
| SEMINARS IN ARTHRITIS AND RHEUMATISM | Modern Rheumatology |
| ARTHRITIS RESEARCH & THERAPY | SCANDINAVIAN JOURNAL OF RHEUMATOLOGY |
| ARTHRITIS CARE & RESEARCH | Archives of Rheumatology |
| JOINT BONE SPINE | ZEITSCHRIFT FÜR RHEUMATOLOGIE |
| Therapeutic Advances in Musculoskeletal Disease | Acta Reumatologica Portuguesa |
| RHEUMATOLOGY INTERNATIONAL | AKTUELLE RHEUMATOLOGIE |
| JOURNAL OF RHEUMATOLOGY | ARP Rheumatology |
| Lupus Science & Medicine |  |

**Supplementary Table 3:**
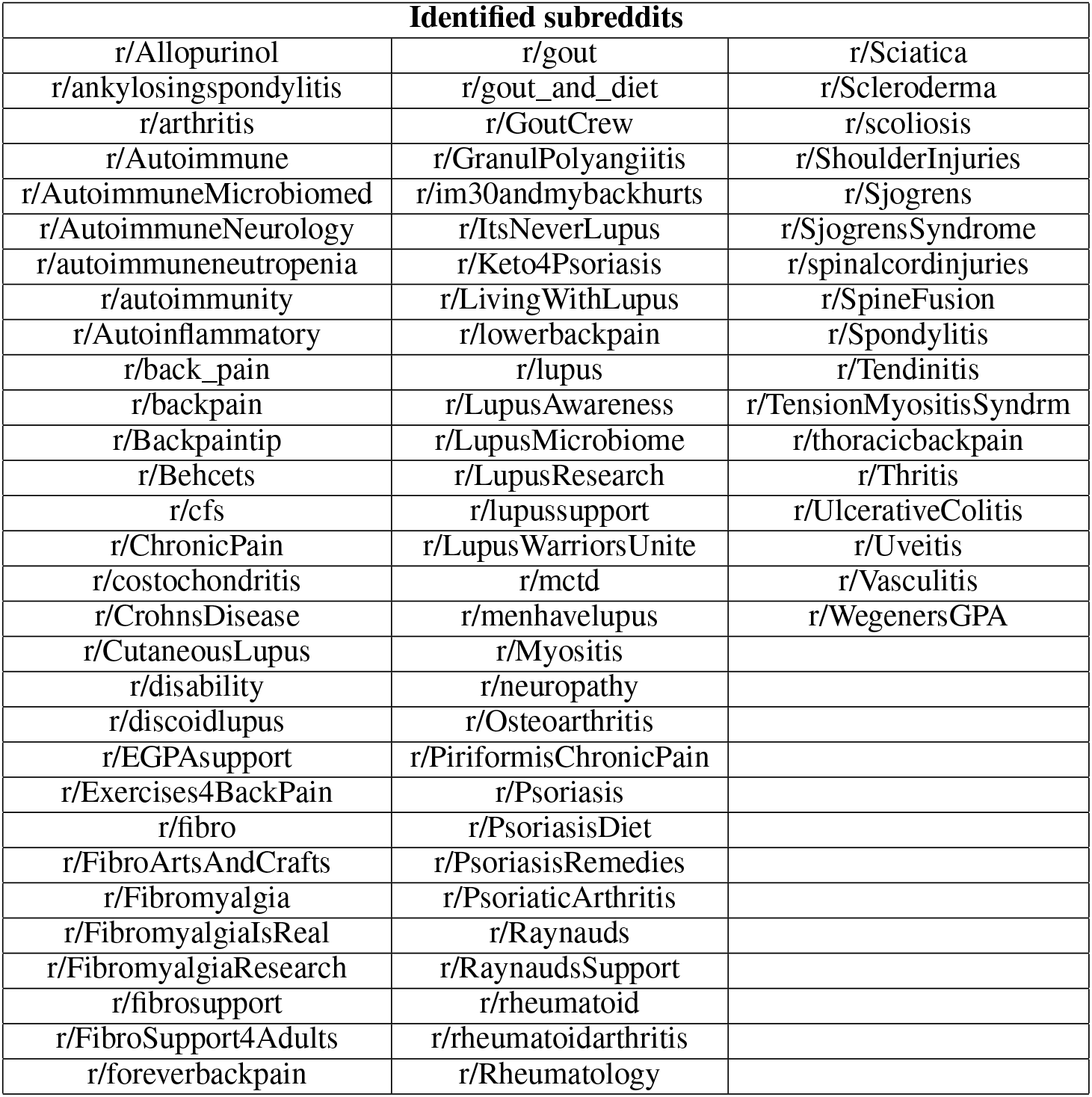
Potential subreddits found through Reddit search engine. Words employed in the search: a*rthritis, autoimmune, back pain, backpain, behcet, behcets, fibromyalgia, gout, lupus, myositis, psoriasis, raynaud, raynauds, rheumatology, scleroderma, sjogren, sjogrens, spondylitis, tendinitis, thritis, uveitis, and vasculitis*

| Identified subreddits |  |  |
| --- | --- | --- |
| r/Allopurinol | r/gout | r/Sciatica |
| r/ankylosingspondylitis | r/gout_and_diet | r/Scleroderma |
| r/arthritis | r/GoutCrew | r/scoliosis |
| r/Autoimmune | r/GranulPolyangiitis | r/ShoulderInjuries |
| r/AutoimmuneMicrobiomed | r/im30andmybackhurts | r/Sjogrens |
| r/AutoimmuneNeurology | r/ItsNeverLupus | r/SjogrensSyndrome |
| r/autoimmuneneutropenia | r/Keto4Psoriasis | r/spinalcordinjuries |
| r/autoimmunity | r/LivingWithLupus | r/SpineFusion |
| r/Autoinflammatory | r/lowerbackpain | r/Spondylitis |
| r/back_pain | r/lupus | r/Tendinitis |
| r/backpain | r/LupusAwareness | r/TensionMyositisSyndrm |
| r/Backpaintip | r/LupusMicrobiome | r/thoracicbackpain |
| r/Behcets | r/LupusResearch | r/Thritis |
| r/cfs | r/lupussupport | r/UlcerativeColitis |
| r/ChronicPain | r/LupusWarriorsUnite | r/Uveitis |
| r/costochondritis | r/mctd | r/Vasculitis |
| r/CrohnsDisease | r/menhavelupus | r/WegenersGPA |
| r/CutaneousLupus | r/Myositis |  |
| r/disability | r/neuropathy |  |
| r/discoidlupus | r/Osteoarthritis |  |
| r/EGPAsupport | r/PiriformisChronicPain |  |
| r/Exercises4BackPain | r/Psoriasis |  |
| r/fibro | r/PsoriasisDiet |  |
| r/FibroArtsAndCrafts | r/PsoriasisRemedies |  |
| r/Fibromyalgia | r/PsoriaticArthritis |  |
| r/FibromyalgiaIsReal | r/Raynauds |  |
| r/FibromyalgiaResearch | r/RaynaudsSupport |  |
| r/fibrosupport | r/rheumatoid |  |
| r/FibroSupport4Adults | r/rheumatoidarthritis |  |
| r/foreverbackpain | r/Rheumatology |  |

**Supplementary Table 4:**
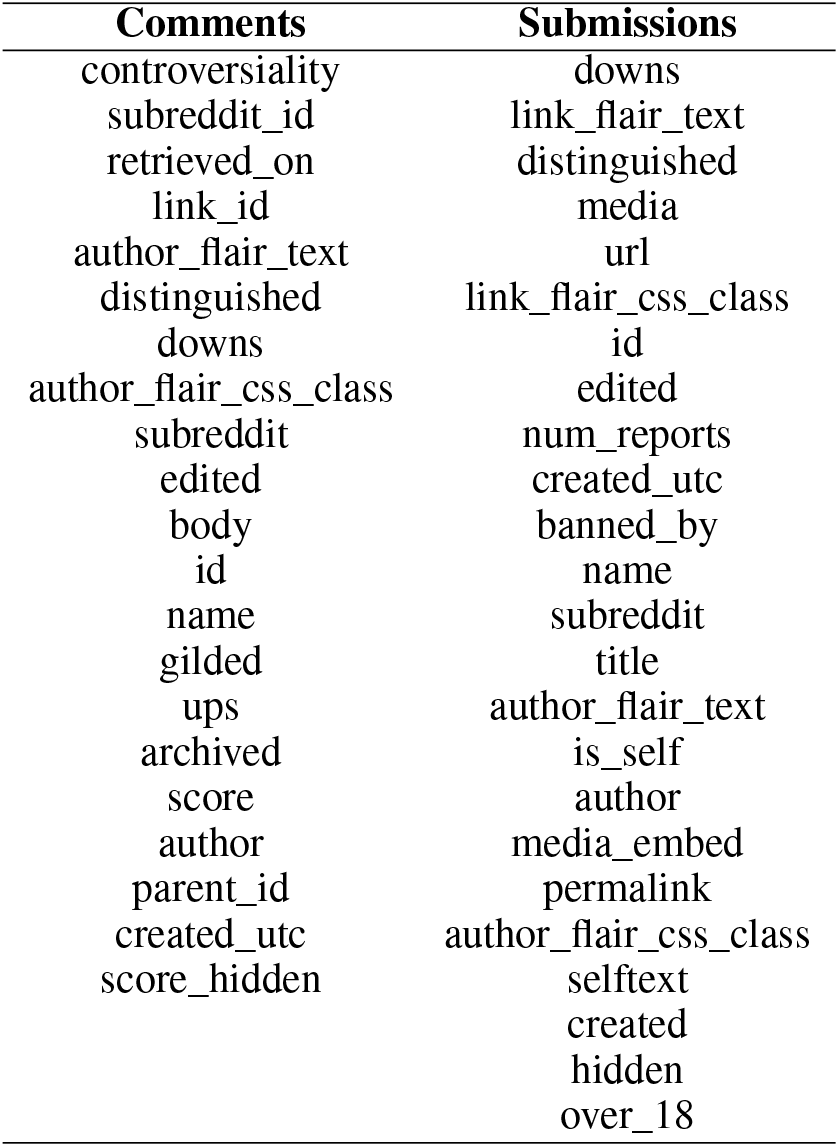
Example of parameters that can be used with *to_csv*.*py* script while decompressing. This is not an exhaustive list

| Comments | Submissions |
| --- | --- |
| controversiality | downs |
| subreddit_id | link_flair_text |
| retrieved_on | distinguished |
| link_id | media |
| author_flair_text | url |
| distinguished | link_flair_css_class |
| downs | id |
| author_flair_css_class | edited |
| subreddit | num_reports |
| edited | created_utc |
| body | banned_by |
| id | name |
| name | subreddit |
| gilded | title |
| ups | author_flair_text |
| archived | is_self |
| score | author |
| author | media_embed |
| parent_id | permalink |
| created_utc | author_flair_css_class |
| score_hidden | selftext |
|  | created |
|  | hidden |
|  | over_18 |

**Supplementary Table 5:** Number of articles with abstract published by year, considering the 34 JCR journals with the category “RHEUMATOLOGY SCIE”. Although 2024 appears, it must be noted that the time interval studied is 2000-2023. This inconsistency is due to the difference in creation and indexing in PubMed and the date of publication.

| Journal | 2000 | 2001 | 2002 | 2003 | 2004 | 2005 | 2006 | 2007 | 2008 | 2009 | 2010 | 2011 | 2012 | 2013 | 2014 |
| --- | --- | --- | --- | --- | --- | --- | --- | --- | --- | --- | --- | --- | --- | --- | --- |
| Acta Reumatol Port | 0 | 0 | 0 | 0 | 0 | 0 | 27 | 38 | 46 | 74 | 60 | 45 | 42 | 34 | 48 |
| Adv Rheumatol | 0 | 0 | 0 | 0 | 0 | 0 | 0 | 0 | 0 | 0 | 0 | 0 | 0 | 0 | 0 |
| Ann Rheum Dis | 158 | 199 | 205 | 219 | 278 | 352 | 302 | 303 | 299 | 295 | 386 | 361 | 320 | 306 | 303 |
| Arch Rheumatol | 0 | 0 | 0 | 0 | 0 | 0 | 0 | 0 | 0 | 0 | 0 | 0 | 0 | 0 | 0 |
| ARP Rheumatol | 0 | 0 | 0 | 0 | 0 | 0 | 0 | 0 | 0 | 0 | 0 | 0 | 0 | 0 | 0 |
| Arthritis Care Res (Hoboken) | 0 | 0 | 0 | 0 | 0 | 0 | 0 | 0 | 0 | 0 | 203 | 188 | 222 | 238 | 226 |
| Arthritis Res Ther | 0 | 0 | 0 | 88 | 108 | 201 | 232 | 181 | 211 | 292 | 308 | 305 | 334 | 284 | 292 |
| Arthritis Rheumatol | 0 | 0 | 0 | 0 | 0 | 0 | 0 | 0 | 0 | 0 | 0 | 0 | 0 | 0 | 332 |
| Best Pract Res Clin Rheumatol | 0 | 51 | 54 | 60 | 55 | 65 | 71 | 65 | 70 | 61 | 68 | 63 | 58 | 57 | 59 |
| BMC Musculoskelet Disord | 1 | 10 | 25 | 28 | 49 | 61 | 103 | 129 | 171 | 167 | 287 | 289 | 265 | 365 | 450 |
| Clin Exp Rheumatol | 150 | 178 | 178 | 190 | 176 | 176 | 167 | 207 | 229 | 248 | 241 | 227 | 252 | 247 | 278 |
| Clin Rheumatol | 107 | 102 | 109 | 104 | 117 | 133 | 198 | 457 | 301 | 251 | 220 | 237 | 249 | 276 | 258 |
| Curr Opin Rheumatol | 82 | 76 | 94 | 98 | 95 | 90 | 86 | 84 | 95 | 88 | 95 | 88 | 91 | 96 | 98 |
| Curr Rheumatol Rep | 62 | 65 | 63 | 57 | 54 | 60 | 59 | 60 | 59 | 58 | 63 | 69 | 80 | 90 | 79 |
| Int J Rheum Dis | 0 | 0 | 0 | 0 | 0 | 0 | 0 | 0 | 0 | 61 | 77 | 59 | 79 | 95 | 110 |
| J Clin Rheumatol | 57 | 63 | 54 | 53 | 65 | 66 | 60 | 67 | 66 | 76 | 78 | 85 | 68 | 71 | 70 |
| J Rheumatol | 427 | 402 | 367 | 402 | 346 | 368 | 371 | 331 | 327 | 365 | 325 | 348 | 304 | 233 | 290 |
| Joint Bone Spine | 83 | 64 | 90 | 81 | 103 | 89 | 121 | 108 | 129 | 122 | 120 | 113 | 101 | 106 | 79 |
| Lancet Rheumatol | 0 | 0 | 0 | 0 | 0 | 0 | 0 | 0 | 0 | 0 | 0 | 0 | 0 | 0 | 0 |
| Lupus | 122 | 137 | 117 | 153 | 157 | 167 | 143 | 143 | 139 | 195 | 218 | 192 | 237 | 201 | 208 |
| Lupus Sci Med | 0 | 0 | 0 | 0 | 0 | 0 | 0 | 0 | 0 | 0 | 0 | 0 | 0 | 0 | 26 |
| Mod Rheumatol | 49 | 65 | 65 | 66 | 91 | 84 | 78 | 100 | 105 | 105 | 105 | 116 | 138 | 193 | 167 |
| Nat Rev Rheumatol | 0 | 0 | 0 | 0 | 0 | 0 | 0 | 0 | 0 | 72 | 83 | 84 | 71 | 82 | 82 |
| Osteoarthritis Cartilage | 73 | 106 | 109 | 96 | 118 | 128 | 168 | 173 | 206 | 216 | 221 | 170 | 194 | 238 | 230 |
| Pediatr Rheumatol Online J | 0 | 0 | 0 | 0 | 0 | 0 | 0 | 22 | 20 | 21 | 30 | 35 | 39 | 46 | 52 |
| Rheum Dis Clin North Am | 56 | 48 | 50 | 46 | 49 | 46 | 44 | 39 | 57 | 55 | 43 | 40 | 49 | 44 | 47 |
| Rheumatol Int | 40 | 70 | 86 | 70 | 78 | 160 | 204 | 182 | 222 | 257 | 240 | 268 | 642 | 458 | 232 |
| Rheumatol Ther | 0 | 0 | 0 | 0 | 0 | 0 | 0 | 0 | 0 | 0 | 0 | 0 | 0 | 0 | 4 |
| Rheumatology (Oxford) | 181 | 174 | 182 | 214 | 226 | 226 | 240 | 292 | 342 | 290 | 289 | 286 | 298 | 276 | 278 |
| RMD Open | 0 | 0 | 0 | 0 | 0 | 0 | 0 | 0 | 0 | 0 | 0 | 0 | 0 | 0 | 0 |
| Scand J Rheumatol | 74 | 67 | 71 | 64 | 73 | 80 | 80 | 69 | 72 | 73 | 75 | 70 | 69 | 78 | 73 |
| Semin Arthritis Rheum | 41 | 43 | 43 | 31 | 37 | 56 | 43 | 44 | 43 | 47 | 50 | 93 | 73 | 84 | 102 |
| Ther Adv Musculoskelet Dis | 0 | 0 | 0 | 0 | 0 | 0 | 0 | 0 | 0 | 9 | 28 | 27 | 33 | 22 | 17 |
| Z Rheumatol | 77 | 34 | 66 | 66 | 45 | 55 | 75 | 77 | 77 | 93 | 96 | 100 | 91 | 88 | 86 |

| Journal | 2015 | 2016 | 2017 | 2018 | 2019 | 2020 | 2021 | 2022 | 2023 | 2024 | Total |
| --- | --- | --- | --- | --- | --- | --- | --- | --- | --- | --- | --- |
| Acta Reumatol Port | 50 | 48 | 40 | 38 | 43 | 41 | 39 | 0 | 0 | 0 | 713 |
| Adv Rheumatol | 0 | 0 | 0 | 40 | 61 | 47 | 71 | 45 | 56 | 0 | 320 |
| Ann Rheum Dis | 311 | 295 | 275 | 225 | 192 | 182 | 171 | 187 | 186 | 0 | 6,310 |
| Arch Rheumatol | 18 | 52 | 77 | 49 | 59 | 73 | 77 | 70 | 22 | 0 | 497 |
| ARP Rheumatol | 0 | 0 | 0 | 0 | 0 | 0 | 0 | 46 | 38 | 0 | 84 |
| Arthritis Care Res (Hoboken) | 201 | 221 | 225 | 222 | 179 | 200 | 209 | 229 | 320 | 27 | 3,110 |
| Arthritis Res Ther | 372 | 288 | 283 | 276 | 295 | 282 | 290 | 269 | 235 | 0 | 5,426 |
| Arthritis Rheumatol | 318 | 283 | 209 | 193 | 197 | 194 | 224 | 181 | 236 | 25 | 2,392 |
| Best Pract Res Clin Rheumatol | 59 | 63 | 63 | 65 | 57 | 57 | 39 | 45 | 56 | 0 | 1,361 |
| BMC Musculoskelet Disord | 386 | 497 | 549 | 455 | 638 | 843 | 1037 | 1130 | 962 | 0 | 8,897 |
| Clin Exp Rheumatol | 285 | 276 | 267 | 265 | 270 | 290 | 283 | 300 | 326 | 21 | 5,727 |
| Clin Rheumatol | 299 | 411 | 368 | 440 | 436 | 443 | 528 | 383 | 336 | 79 | 6,842 |
| Curr Opin Rheumatol | 83 | 91 | 91 | 89 | 91 | 78 | 74 | 50 | 54 | 20 | 2,077 |
| Curr Rheumatol Rep | 77 | 75 | 81 | 86 | 76 | 90 | 76 | 41 | 31 | 5 | 1,616 |
| Int J Rheum Dis | 105 | 159 | 224 | 269 | 274 | 186 | 158 | 155 | 277 | 58 | 2,346 |
| J Clin Rheumatol | 60 | 50 | 51 | 49 | 61 | 68 | 130 | 163 | 83 | 11 | 1,725 |
| J Rheumatol | 301 | 264 | 241 | 188 | 195 | 200 | 228 | 166 | 232 | 0 | 7,221 |
| Joint Bone Spine | 75 | 96 | 92 | 87 | 84 | 70 | 86 | 79 | 92 | 8 | 2,278 |
| Lancet Rheumatol | 0 | 0 | 0 | 0 | 23 | 63 | 65 | 64 | 57 | 5 | 277 |
| Lupus | 196 | 197 | 202 | 281 | 212 | 210 | 262 | 200 | 178 | 19 | 4,486 |
| Lupus Sci Med | 29 | 35 | 27 | 45 | 39 | 46 | 73 | 89 | 70 | 0 | 479 |
| Mod Rheumatol | 162 | 161 | 167 | 151 | 145 | 144 | 159 | 156 | 265 | 0 | 3,037 |
| Nat Rev Rheumatol | 82 | 63 | 62 | 53 | 51 | 54 | 53 | 55 | 58 | 4 | 1,009 |
| Osteoarthritis Cartilage | 255 | 239 | 241 | 188 | 194 | 153 | 166 | 144 | 164 | 20 | 4,210 |
| Pediatr Rheumatol Online J | 61 | 67 | 80 | 82 | 83 | 90 | 162 | 109 | 138 | 0 | 1,137 |
| Rheum Dis Clin North Am | 44 | 46 | 44 | 45 | 43 | 51 | 48 | 56 | 54 | 13 | 1,157 |
| Rheumatol Int | 254 | 207 | 245 | 310 | 244 | 234 | 237 | 235 | 270 | 39 | 5,484 |
| Rheumatol Ther | 15 | 25 | 37 | 44 | 46 | 66 | 127 | 97 | 110 | 11 | 582 |
| Rheumatology (Oxford) | 267 | 262 | 264 | 282 | 261 | 445 | 666 | 518 | 686 | 0 | 7,445 |
| RMD Open | 80 | 86 | 112 | 113 | 107 | 128 | 147 | 205 | 270 | 0 | 1,248 |
| Scand J Rheumatol | 72 | 74 | 62 | 58 | 59 | 58 | 60 | 56 | 79 | 9 | 1,675 |
| Semin Arthritis Rheum | 103 | 107 | 116 | 125 | 155 | 204 | 165 | 150 | 171 | 27 | 2,153 |
| Ther Adv Musculoskelet Dis | 22 | 18 | 27 | 22 | 31 | 81 | 107 | 107 | 48 | 0 | 599 |
| Z Rheumatol | 94 | 97 | 105 | 95 | 101 | 116 | 105 | 106 | 132 | 7 | 2,084 |

**Supplementary Table 6:** Submissions, comments and ratio comments/submissions for each subreddit

| Reddit community | Submissions<br>(S) | Comments<br>(C) | Ratio C/S |
| --- | --- | --- | --- |
| r/ankylosingspondylitis | 16820 | 189710 | 11.28 |
| r/Autoimmune | 6356 | 39893 | 6.28 |
| r/autoimmunity | 5938 | 36400 | 6.13 |
| r/backpain | 29519 | 139770 | 4.73 |
| r/Fibromyalgia | 62516 | 716859 | 11.47 |
| r/gout | 15395 | 170018 | 11.04 |
| r/lupus | 30553 | 270520 | 8.85 |
| r/PsoriaticArthritis | 8821 | 105519 | 11.96 |
| r/rheumatoid | 18135 | 193709 | 10.68 |
| r/rheumatoidarthritis | 6168 | 58170 | 9.43 |
| r/Sjogrens | 6377 | 71941 | 11.28 |
| r/Thritis | 8464 | 61740 | 7.29 |

### Supplementary Figures

**Supplementary Figure 1:**
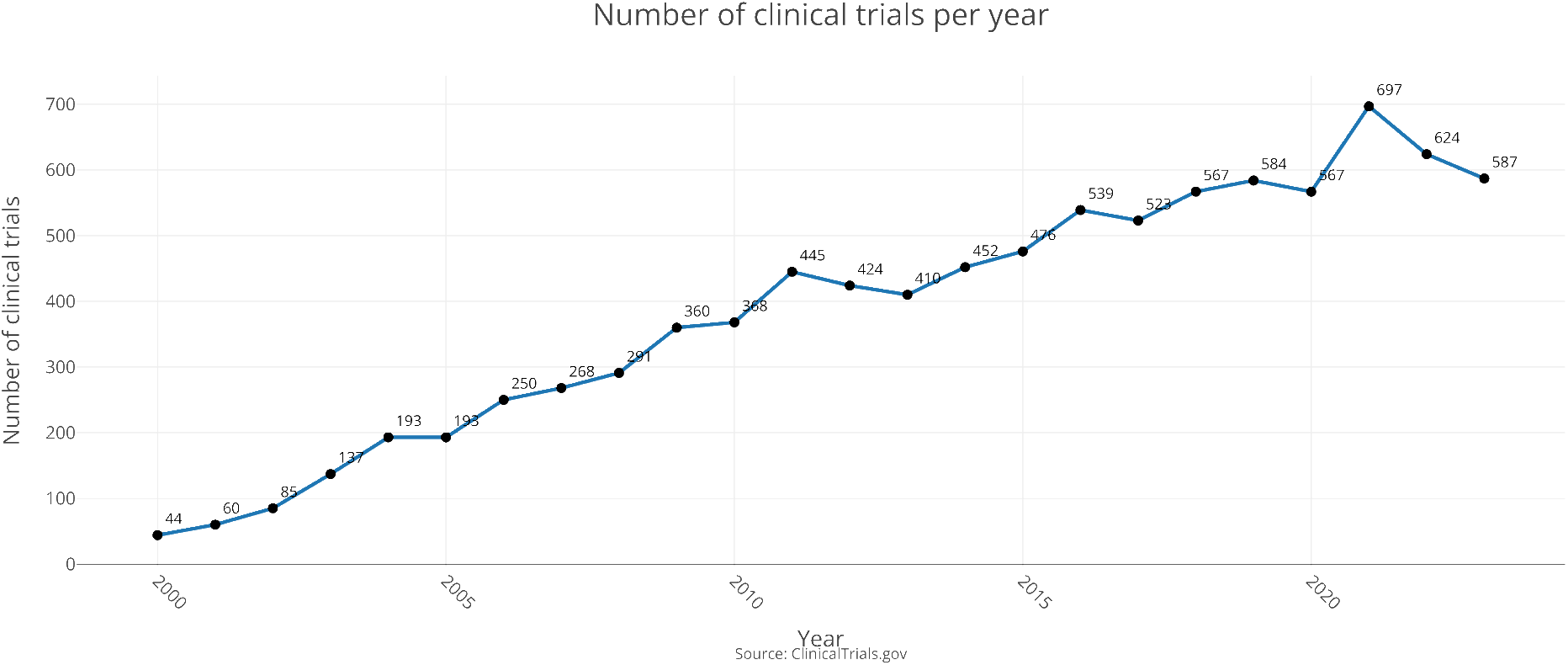
RMDs related clinical trials per year. Only data up to 2023 is shown

